# Predictors of intention to use mobile health apps for comprehensive sexuality education among young people in the Democratic Republic of Congo: a correlational study

**DOI:** 10.64898/2026.04.09.26350561

**Authors:** François Kajiramugabi Maneraguha, José Côté, Anne Bourbonnais, Caroline Arbour, Miguel Chagnon, Marie Hatem

## Abstract

**Background:** Comprehensive sexuality education (CSE) is essential to the health and well-being of young people. In the Democratic Republic of Congo (DRC), where more than 65% of the population is under the age of 25, access to interpersonal CSE remains limited owing to sociocultural and structural barriers. This exposes young people to persistent socio-sanitary vulnerabilities. In this context, mobile health apps (MHAs) constitute a promising solution, supported by the growing use of smartphones among young Congolese. However, this group’s intention to use MHAs for CSE has been the subject of little research to date.

**Objective:** The aim of this study was to identify predictors of intention to use MHAs among young Congolese, based on the extended Unified Theory of Acceptance and Use of Technology (UTAUT2).

**Methods:** A predictive correlational study was conducted in eight public secondary schools in Bukavu (DRC) with a stratified random sample of 859 students. Predictors of intention to use—performance expectancy (PE), effort expectancy (EE), social influence (SI), facilitating conditions (FC), and perceived risk (PR)—and moderators—age, gender, and past MHA experience—were measured from data collected through a self-administered UTAUT questionnaire. Descriptive and multivariate analyses were run on SPSS version 28.

**Results:** Mean age of participants was 16.3 years (*SD* = 1.5). Boys made up 55.1% of the sample. Overall, 51.0% of the sample owned a smartphone, of which 62.3% reported having easy access to mobile data and 16.2% were already using MHAs to learn about sexual health. Intention to use MHAs was positively influenced by PE (*β* = 0.523, *p* < 0.001), EE (*β* = 0.115, *p* < 0.001), and SI (*β* = 0.113, *p* < 0.001). FC (*p* = 0.260) and PR (*p* = 0.631), however, had no significant influence. Age moderated all of the relationships tested (*F* (1, 849–854) = 9.97–20.82; *p* ≤ 0.002), with more marked effects observed among younger participants 14–15 years old. The final model explained 44% of the variance, indicating good predictive power.

**Conclusion:** Intention to use digital CSE was explained primarily by PE, EE, and SI and moderated by age. To strengthen this intention, stakeholders will need to promote e-interventions that are pertinent, easy to use, socially valorized, and tailored to young people’s needs and to the local context.

**Author Summary:** Young people in the Democratic Republic of Congo often face barriers to accessing reliable comprehensive sexuality education, including social taboos and limited resources. At the same time, smartphones are becoming more common, creating new opportunities to deliver this information through mobile health applications. In this study, we explored what drives young people’s willingness to use mobile health applications for comprehensive sexuality education. We surveyed 859 students from eight public secondary schools in Bukavu, South Kivu Province, in the eastern Democratic Republic of Congo. We found that young people were more likely to intend to use these applications when they perceived them as useful, easy to use, and socially supported. Concerns about risks, such as privacy, did not reduce their intention. Our findings suggest that mobile health applications could help expand access to comprehensive sexuality education if they are codesigned with young people, tailored to their needs, easy to use, and supported by their social environment. This work highlights the importance of understanding young people’s perspectives before developing digital tools for comprehensive sexuality education, especially in settings where access remains limited.

## 1. INTRODUCTION

Mobile health apps (MHAs) today constitute powerful levers of prevention, positive behavioural change, and promotion for the sexual health of young people. They offer this population continuous, interactive, and confidential access to reliable information and sexual health services (1). MHAs can be defined as programs designed to work on mobile devices (e.g., smartphones, tablets) characterized by small size and low resource consumption. There are presently over 100,000 MHAs available on Android and iOS, and the number is ever-growing as technologies and health education needs evolve (2).

This expansion has occurred alongside the massive proliferation of smartphones, which have become increasingly accessible to young people in the low-income countries of sub-Saharan Africa (3). In Ghana, 88% of young people own a smartphone (4); in Kenya, 90% do (5). Young people thus constitute the category most connected. They use these devices on a daily basis for entertainment, communication, and health information (4,5), thus developing active use behaviours that strengthen their sense of social connection (6). Often given by their parents as a birthday gift or academic performance reward, smartphones are perceived by many young people as tools for socialization, security, empowerment, and social assertion (7).

The near-permanent presence of smartphones and mobile apps in the hands of young people affords a strategic opportunity to bolster their comprehensive sexuality education (CSE). This digital approach is particularly relevant in contexts where access to quality interpersonal CSE remains limited (8) owing to persistent taboos around CSE, stigmatization of sexual health querying, gaps in teacher and caregiver training, and lack of adequate pedagogical material, as is the case in the Democratic Republic of Congo (DRC) (9–12). Numerous MHAs, such as Girl Talk (13), Mosexy (14), Pulse (15), and Tumaini (16,17), were developed in response to these structural and sociocultural barriers. These apps help prevent teenage pregnancies and sexually transmitted infections, encourage use of condoms and contraception services, and contribute to improve knowledge of sexual health. The growing interest among young people attests to the appeal and strong potential acceptability of these apps (18).

However, the degree to which young people engage with these apps diminishes over time, resulting in high rates of rejection (19) or dropout (20) and in reduced effectiveness (18). This situation often stems from the absence of analysis of intention to use prior to designing the apps (20). Two umbrella reviews (1,21) and eight recent systematic reviews (18,22–28) demonstrated that, despite the boom in MHA-driven digital CSE, very few studies have examined the factors associated with young people’s intention to use these apps, especially in Francophone sub-Saharan Africa. Most of the works reviewed have been conducted in Anglophone or high-income contexts, where the countries of Francophone sub-Saharan Africa, including the DRC, have figured very little (1,21,25). Moreover, the studies have focused more on past MHA use than on intention to use. This gap is critical, as it limits the transferability of results to Francophone contexts. The lack of transferability is attributable to language, but also to marked differences in the access young people have to formal secondary education, in the political and institutional organization of the CSE offering, in the cultural norms governing parent-child exchanges regarding sexuality (29,30), and in the uneven reach of mobile Internet service and technological innovation—all factors and phenomena particularly salient in Francophone sub-Saharan Africa (31,32). From this perspective, it is indispensable to analyze young people’s intention to use apps for CSE. Kim and Park (33) underscored fundamental importance of analyzing this intention beforehand, as it determined how effective app use would be and what impact e-interventions could have on the health of a population. Intention to use has been defined as the extent to which a user expects to use mobile health regularly in the future (34). Identifying the factors that influence this is essential to designing CSE MHAs that are acceptable, effective, and durable and that thus deliver high value to the target population. The central role played by intention has been confirmed by various primary and secondary studies recently (35,36).

A systematic review (35) found that 68 factors influenced MHA acceptability and that intention to use was the most determinant of these. The study found, also, that intention was influenced, in turn, by perceived usefulness, ease of use, social influence, perceived risk for confidentiality, and attitude towards the tool, underscoring the importance of analyzing these dimensions prior to designing and implementing digital tools of the sort. Similarly, a meta-analysis that covered 35 empirical studies (36) showed that perceived usefulness, ease of use, trust, and perceived risk significantly influenced intention to use MHAs. These factors were observed among young people in a variety of contexts, including the promotion of physical activity and nutrition, reproductive health, sexuality education, and access to information on sexuality via social media.

In Europe, studies have evidenced the central role played by individual and motivational factors in the intention of young people to use MHAs. In Germany, Van de Werken et al. (34) demonstrated, in a study grounded in the Unified Theory of Acceptance and Use of Technology (UTAUT), that 82.5% of intention to use MHAs among 242 users ages 18 and over was predicted by five main factors: compatibility, habit, performance expectancy, age, and price value. For their part, Bickmann et al. (37) confirmed that performance expectancy and hedonic motivation influenced intention to use educational MHAs. However, the fact that social influence remained weak in this study illustrated the need for social features better adapted to young users. In Sweden, Altmann and Gries (38) identified perceived usefulness and available time as the principal factors associated with intention to use MHAs among 102 urban youths 24 to 29 years old recruited by way of purposive and snowball sampling. In Poland, Burzyńska et al. (39) reported that performance expectancy, habit, and place of residence were its strongest predictors. Concerns about confidentiality appeared to be the weakest factor, and age, gender, education level, and type of academic establishment had no notable influence. Overall, these results suggested that, in the European context, perceived effectiveness and technological familiarity were more important than sociodemographic characteristics in young people’s intention to use MHAs.

In Asia, the literature on this intention has been more mixed. In China, in their study based on the Technology Acceptance Model (TAM) and Innovation Resistance Theory, which involved 879 young university students for the most part 18 to 20 years old, Yang et al. (40) showed that attitude towards MHAs was the principal predictor of intention to use, followed by perceived usefulness, ease of use, perceived risk, complexity, and barriers related to perceived value. For their part, in a cross-sectional study based on the TAM, To et al. (41) underscored that perceived usefulness—bolstered by effective communication and a heightened health awareness—significantly influenced this intention among 468 young people 20 to 39 years old. In Malaysia, Yasin et al. (42) reported, in a UTAUT-based correlational study, that intention to use MHAs among 312 young people 23 to 27 years old was influenced by performance expectancy, facilitating conditions, and health awareness, whereas effort expectancy and social influence did not prove significant. In Korea, Park et al. (43) observed, in an online survey based on the TAM and conducted with a convenience sample of 329 MHA users 18 years of age, that ease of use, perceived usefulness, and user satisfaction reinforced intention to use. Together, these results confirmed, through the different theoretical models mobilized in the Asian context, the primacy of perceived usefulness, ease of use, and user satisfaction in predicting intention among young MHA users. However, most of the studies reviewed focused on young people 18 and over, primarily from university or extra-academic settings. This state of knowledge accentuated the pertinence of first examining intention to use MHAs at an earlier stage of the life course, when needs regarding the prevention of pregnancy and infection, the promotion of responsible sexual behaviour, and the supply of educational support are strongest.

Few studies have explored these dynamics in Francophone sub-Saharan Africa, on which the literature remains very limited. A few studies have been conducted in Anglophone countries, especially Kenya, Nigeria, and South Africa, but they have looked at intention among young people to use digital tools in general and not specifically MHAs for CSE. One of these (44) showed that performance expectancy, social influence, effort expectancy, facilitating conditions, and attitude were significantly associated with intention among 936 young people 18 to 24 years old to access CSE through the social networks. In rural Kenyan settings, another study (45) revealed that perceived usefulness and attitude had a positive influence among young people 18 to 35 on intention to use a digital tool for their CSE. In Francophone sub-Saharan Africa, no empirical study to date has, to our knowledge, analyzed the predictors of intention to use MHAs for CSE among young people 15 to 24 years of age. In the DRC, the rare works available have concerned limited e-interventions, such as by way of radio texting (46) and chatbots (47), but these did not analyze intention to use.

Against this background, we undertook a study to examine intention to use MHAs for CSE among young people in the DRC, a country where 15- to 24-year-olds make up more than 65% of the population and where digital CSE constitutes a promising, innovative way to improve their access to information that is reliable, confidential and tailored to their needs (48). Our aim was to identify the predictors of this intention among young people in Bukavu, based on the extended UTAUT (UTAUT2).

Initially developed by Venkatesh et al. (49), the UTAUT constitutes one of the go-to theories for predicting technology use and acceptance, on account of its high predictive and explanatory power (50). It was extended to apply to mobile health interventions over the past two decades and has demonstrated high predictive power for explaining the barriers and facilitators of intention to use these interventions, including among young people (34,51). In this extended version, referred to as UTAUT2, Venkatesh et al. (52) identified seven predictors of behavioural intention (BI) to use technology, as shown in Fig 1 : i) performance expectancy (PE) corresponds to perceived usefulness; ii) effort expectancy (EE) corresponds to ease of use ; iii) social influence (SI) corresponds to expected social support; iv) facilitating conditions (FC) refer to the availability of infrastructure to support use; v) habit (H) refers to the individual’s tendency to use MHAs on a regular basis; vi) hedonic motivation (HM) refers to pleasure or satisfaction expected from use; and vii) price value (PV) refers to the degree to which users estimate that the MHA is worth the asking price or the effort required to use it. Under this model, these predictors are moderated by age, gender, and past experience with the technology (34,49,52).

**Fig 1:**
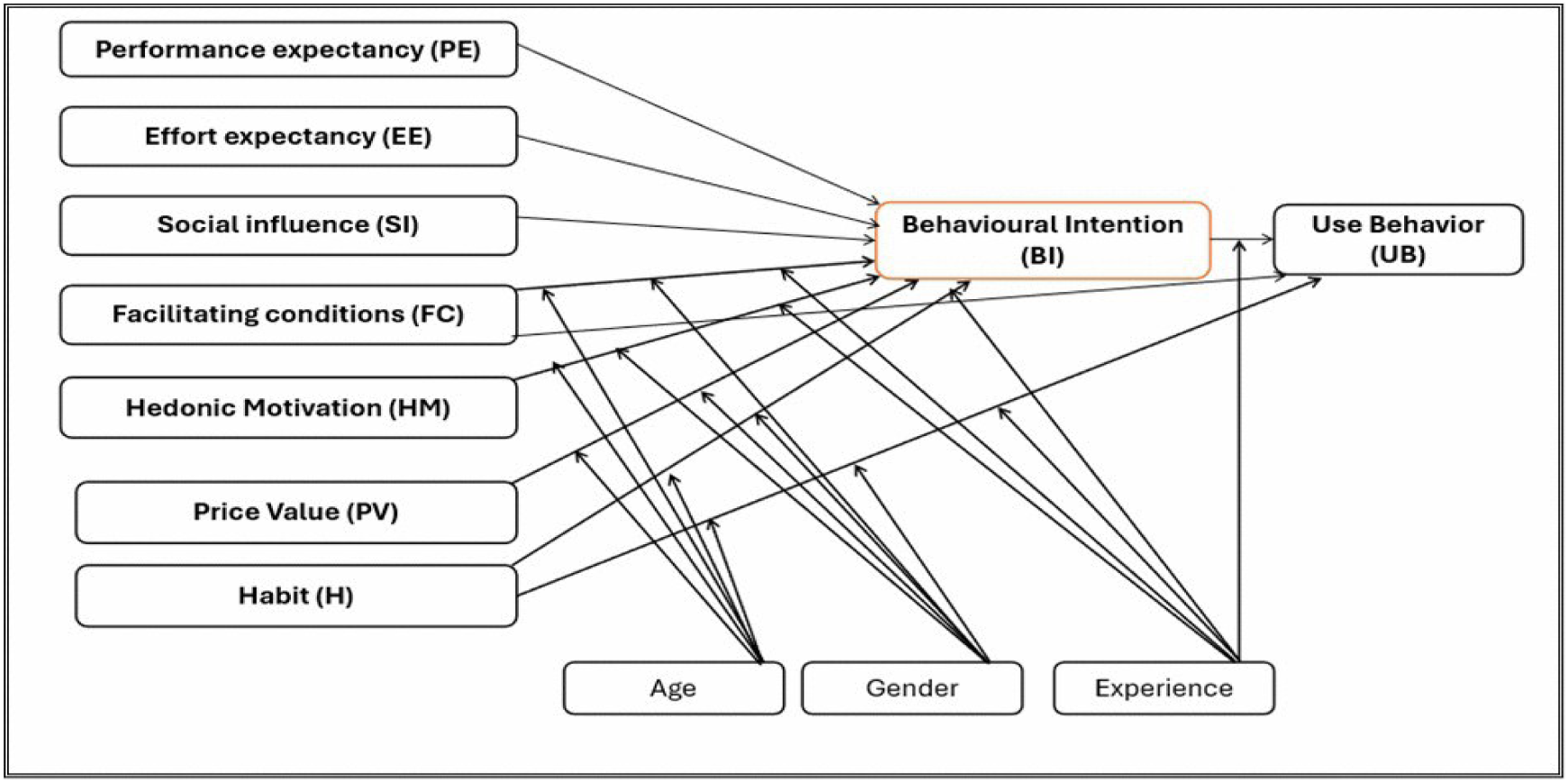
Frame of reference (UTAUT2): predictors and moderators of behavioural intention to use technology and use behaviour *(52, p.160)*

For the purposes of our study, four core predictors of BI from the UTAUT2—PE, EE, SI, and FC—and the moderators—age, gender, and experience—were selected. Following recommendations to adapt the UTAUT to the specific context where it is to be used (53), the model was enhanced by adding perceived risk (PR) as a contextual factor. This was defined as the perception of the uncertainty and the gravity of potential consequences regarding the use of an MHA, including anxiety related to the Internet and the perception that the digital environment is threatening or unsafe and acts as a damper on intention to use (34,54). This approach fits within a double logic: application of the UTAUT in a specific context (*UTAUT in context*) and an analysis of the effects of contextual constraints on the mechanisms of acceptance (*UTAUT of context*) (53,55), by examining how these factors modulate the relationships between the model’s variables and BI. On the other hand, HM, PV, and H were not retained as they are associated more with prolonged use behaviour (56) and are less relevant in developing countries where digital CSE is emerging and where utilitarian factors have prevalence over other factors (57). The variables selected (PE, EE, SI, FC, PR, age, gender, and experience) thus allowed us to incorporate both the individual and contextual dimensions of young people’s intention to use MHAs for CSE, as illustrated in Fig 2.

**Figure 2:**
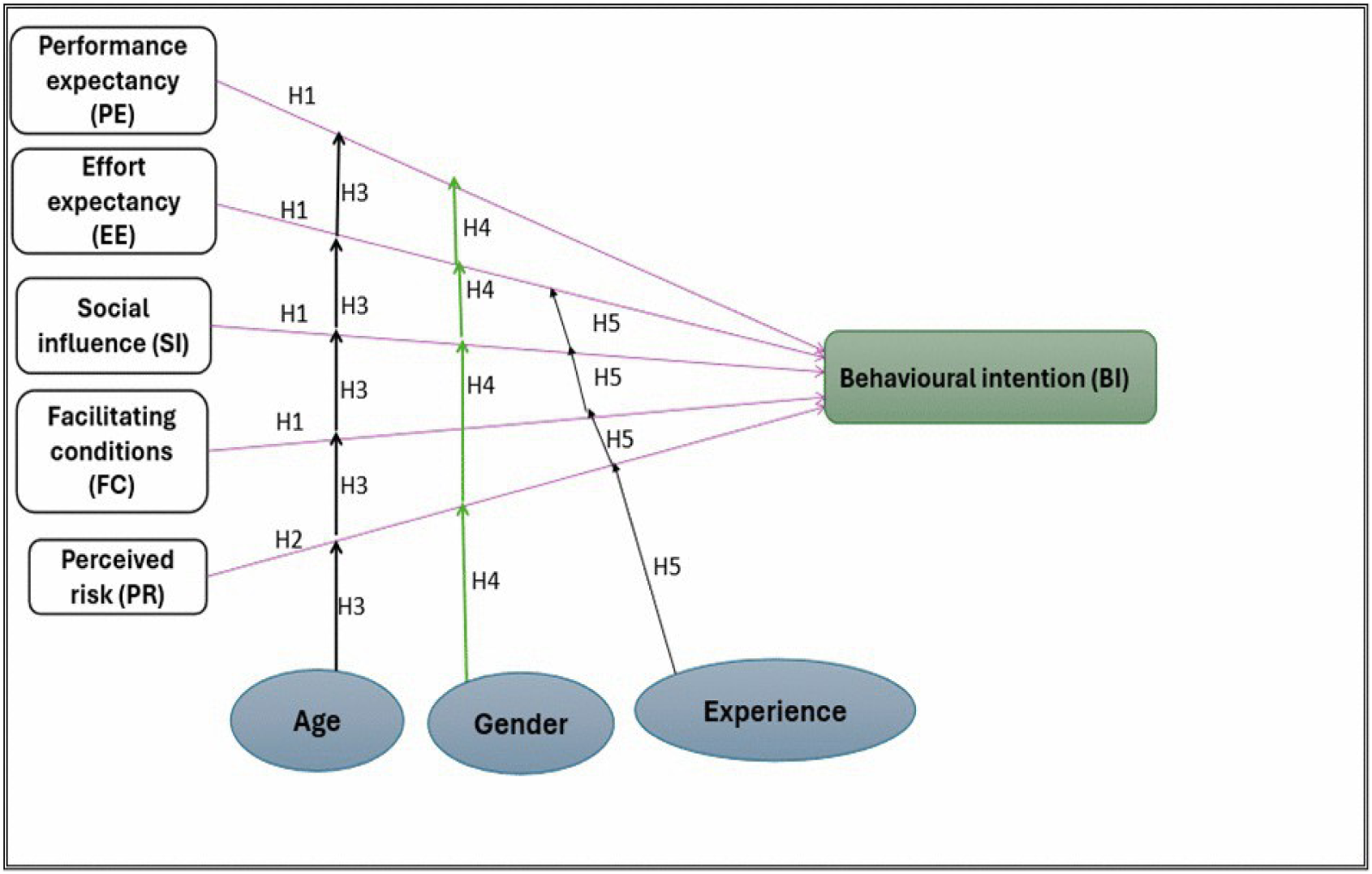
Research hypotheses regarding intention to use MHAs. On this basis, we formulated five hypotheses: **H1**: PE, EE, SI, and FC would prove to have a positive influence on the intention of young people in the DRC to use MHAs for their CSE. **H2**: PR would have a negative influence on intention to use MHAs for CSE. **H3**: Age would have a moderating effect on the relationships between PE, EE, SI, FC, PR and intention to use, and the effect would be differentiated by age. **H4**: Gender would moderate the relationships between PE, EE, SI, FC, PR and intention to use MHAs for CSE. **H5**: Past MHA experience would have a positive moderating effect on the relationships between PE, EE, SI, FC, and PR relative to intention to use.

## 2. METHOD

### 2.1. Design

We undertook a predictive, correlational study with an observational, cross-sectional design (58). The various methodological elements are presented below following the STROBE (Strengthening the Reporting of Observational Studies in Epidemiology) guidelines (59,60). For more information, see S2 Appendix.

### 2.2. Setting

The study was carried out in the fall of 2024 in eight public secondary schools in Bukavu, Sud-Kivu province in the eastern part of the DRC, across three communes (Bagira, Ibanda, and Kadutu) and four educational zones. The participating establishments included five confessional schools—three catholic (Collège Alfajiri, Institut Avenir, Institut Kasali) and two protestant (Institut Bangu, Institut Imani Panzi)—and three state schools (Institut de Kadutu, Athénée d’Ibanda, Institut de Bagira).

### 2.3. Participants and inclusion criteria

Participants were students 15 to 24 years old enrolled at the selected schools. This age bracket corresponded to “youth” as defined by the United Nations (61,62) and took account of the local educational context, where students over 18 can still attend secondary school (63). To be included, students had to be on the 2024-2025 school register, be in attendance on the days the survey took place and provide free and informed consent in the case of adults or assent in the case of minors, along with prior consent from school principals in their capacity of institutional legal tutors. Students absent on the day of the survey, or the following day were excluded. Participants were selected by way of stratified random sampling (58) by class, program enrolled in, and school, for a total of 123 strata. Eight students were randomly selected per stratum from the attendance roll using codes generated by the Centre d’intégration et d’analyse de données médicales du Centre Hospitalier de l’Université de Montréal.

### 2.4. Variables

Behavioural intention (BI) to use MHAs for CSE was the dependent variable. The independent variables were PE, EE, SI, FC, and PR. The three moderators were age, gender, and past MHA experience. Other variables were academic and technological in nature. The academic variables regarded the school attended. The technological variables regarded ownership or not of a smartphone and ease of access to mobile data. These variables were used for descriptive purposes to characterize the academic and technological context of the participants and to document the conditions regarding access to MHAs, without their being incorporated as explanatory variables in the correlational analyses.

### 2.5. Data sources and methods of measurement

The data were collected through the UTAUT questionnaire, an instrument developed by Venkatesh et al. (49) to analyze intention to use technology and its predictors. For our study, we used the French version validated by Baudier et al. (64). This version was shown to possess robust psychometric properties: high internal consistency, with Cronbach’s alpha and composite reliability values above 0.70 for all the subscales; acceptable convergent validity, with Average Variance Extracted (AVE) values above 0.50; and divergent validity confirmed by the fact that the square root of AVE has proved greater than all inter-construct correlations. Moreover, the overall quality of the model has been deemed satisfactory (goodness-of-fit = 0.71) and its predictive power has been shown to be high (*R*² = 0.712) regarding intention to use (64).

The hardcopy questionnaire was self-administered in a school setting that would ensure confidentiality. It comprised two sections. The first served to collect sociodemographic, academic and technological data; the second measured intention to use MHAs for CSE and its predictors, through items formulated specifically for CSE. The dependent variable (BI) was measured through three items. The independent variables were each measured by way of four items, except for PR, which had only three. In all, the questionnaire comprised 22 items rated on a five-point Likert scale ranging from “strongly disagree” to “strongly agree” distributed across six subscales corresponding to the variables under study. The items and subscales are detailed in S1 Appendix. The questionnaire was pre-tested with 15 students from two public secondary schools not included in the main sample, one state and one confessional, to ensure clarity of wording and ease of response. Students completed the questionnaire in 15 to 20 minutes. Internal consistency was assessed through Cronbach’s alpha. Values obtained ranged from 0.539 to 0.727: BI (α = 0.727), PE (α = 0.674), EE (α = 0.657), SI (α = 0.593), FC (α = 0.539), and PR (α = 0.563). Though some subscales scored below the recommended 0.70 threshold (65), they could nevertheless be deemed acceptable in an explanatory context, especially in light of the small number of items in each subscale. According to Hair et al. (66), values from 0.60 to 0.70 are considered adequate for scales comprising a small number of items, as was the case in our study (3 or 4 items per subscale).

### 2.6. Bias and mitigation strategies

Our study could potentially be affected by selection bias, social desirability bias, and missing data. To limit selection bias, the data were collected from students present the day of the survey or the day after, following a schedule coordinated with school administrators. To attenuate social desirability bias we anonymized the questionnaire and had it distributed by independent researchers trained in the research objectives and in the data collection instrument. These researchers, who had backgrounds in nursing science, public health, and sociology, took part in simulation exercises to harmonize administration of the data collection instruments and to ensure data quality. None of the selected student refused to participate in the study and no questionnaire was returned incomplete. To reduce missing data bias, we checked that all questionnaires were completed in full and that responses were consistent.

### 2.7. Sample size

Sample size was estimated on the basis of the classic rule recommended for multiple linear regression analyses, namely, a minimum of ten observations per independent variable (67). Given the 11 variables included in the model, a sample of 110 participants was required to ensure appropriate statistical power. However, to take account of the stratification, the heterogeneity of academic contexts, and a conservative planning approach, this figure was multiplied by the number of survey sites (n = 8 secondary schools), which set the target at 880 students. In the end, 859 students were surveyed, as 21 were absent on the days the survey was held.

### 2.8. Statistical analyses

The main quantitative variables corresponded to the mean scores on the subscales: BI, PE, EE, SI, FC, and PR. Participant age was treated as a continuous variable. The other variables, such as gender, smartphone ownership, and school attended, were coded as categorical. Analyses were run on IBM/SPSS version 28 to examine the relationships between the sociodemographic variables, the predictors, and BI. Descriptive analyses (means, standard deviations, frequencies, percentages) allowed us to draw a general portrait of the sample. Simple linear regressions were run to estimate the bivariate associations between each predictor (PE, EE, SI, FC, PR) and BI, as measured by the unstandardized coefficient (*B*) and the standardized coefficient (*β*). Moderating effects were tested and partial eta squared (η*_p_*²) was computed as a measure of effect size. In keeping with the methodological recommendations out of the recent literature (68,69), particularly from one systematic review (70), our study’s data had a hierarchical structure, with students (level 1) nested in schools (level 2). This organization could generate a cluster effect stemming from an intra-class correlation between students attending the same school, having to do with shared institutional factors that were not measured. The potential impact of this structure of dependent observations could result in an under-estimation of standard errors and thus undermine the validity of statistical inferences. Consequently, sensitivity analyses were carried out to document the potential impact on results of the school cluster effect. For these sensitivity analyses, the data were grouped by school to model the potential dependence of the observations.

For the multivariate analyses, we ran linear regressions and mixed linear models with the random school effect. Linearity, normality of residuals, and multicollinearity were checked and found to be within acceptable limits. The variables were centred to test the interactions with age, gender, and past MHA experience. The statistical significance of variables and interactions was assessed by way of *t*-tests and the *F* statistic of the variance analysis table. We computed *p*-value and η*_p_*² for effect size, to measure the portion of the variance explained by each predictor in isolation. The significance threshold was set at alpha = 0.05. Finally, a two-step hierarchical regression model assessed the effect of the significant interactions on change in the coefficient of determination (Δ*R*²) between the two models, by way of the likelihood ratio test. There were no missing data in the sample.

### 2.9. Ethical considerations

The study was approved by the Université de Montréal Health and Science Research Ethics Committee (CERSSES 2024-6039) and by the DRC National Health Ethics Committee (CNES 001/DPSK/222PM/2024). No nominative information was collected. Participants were selected using attendance roll numbers; no personal identifiers were collected and no link to the students’ identity was kept. Participation in the study was entirely voluntary. Participants were advised that they could withdraw from the study at any time without any negative academic consequences. The purpose of the study was clearly explained to them. They were invited to participate, were given the chance to ask all their questions, and they then gave their written consent. Free, informed consent was sought directly from students 18 and over through an appropriate information and consent form. In accordance with Congolese law, consent for minors (15 to 17 years old) was obtained from the school principals acting as their institutional legal tutors. The assent of these students was then sought and obtained through an information form that they signed and remitted prior to data collection.

## 3. RESULTS

### 3.1. Participants’ profile

Table 1 presents the participants’ characteristics. A total of 859 students were included in the study. Mean age was 16 years (±1.54); 55.1% were male. A relative majority of the students were in their third year of secondary school, equivalent to grade 11 (27.2%). Regarding technology, 51% owned a smartphone. Of these, 62.3% reported having easy access to mobile data and 16.2% were already using an MHA to learn about sexual health and had been doing so for about five months on average (±4.83).

**Table 1:** Sociodemographic, academic and technological characteristics of participants.

| Variables |  | School |  |  |  |  |  |  |  |  |
| --- | --- | --- | --- | --- | --- | --- | --- | --- | --- | --- |
|  |  | Institut de Bagira<br>(*n* = 137) | Institut Bangu<br>(*n* = 152) | Athénée d'Ibanda<br>(*n* = 71) | Collège Alfajiri<br>(*n* = 120) | Institut de Kadutu<br>(*n* = 83) | Institut Kasali<br>(*n* = 83) | Institut Avenir<br>(*n* = 124) | Institut Imani Panzi<br>(*n* = 89) | Total<br>(*n* = 859) |
| Gender | Male | 81 (59.1%) | 81 (53.3%) | 37 (52.1%) | 60 (50.0%) | 44 (53.0%) | 50 (60.2%) | 73 (58.9%) | 47 (52.8%) | 473<br>(55.1%) |
|  | Female | 56 (40.9%) | 71 (46.7%) | 34 (47.9%) | 60 (50.0%) | 39 (47.0%) | 33 (39.8%) | 51 (41.1%) | 42 (47.2%) | 386<br>(44.9%) |
| Mean age, in years |  | 16.8±1.66 | 16.58±1.63 | 16.63±1.59 | 15.58±1.2 | 16.45±1.48 | 16.22±1.33 | 16.02±1.51 | 16.34±1.37 | 16.33±1.54 |
| Grade | 1<sup>st</sup> year | 36 (26.3%) | 36 (23.7%) | 9 (12.7%) | 37 (30.8%) | 23 (27.7%) | 20 (24.1%) | 29 (23.4%) | 20 (22.5%) | 210<br>(24.4%) |
|  | 2<sup>nd</sup> year | 38 (27.7%) | 38 (25.0%) | 28 (39.4%) | 11 (9.2%) | 16 (19.3%) | 26 (31.3%) | 31 (25.0%) | 21 (23.6%) | 209<br>(24.3%) |
|  | 3<sup>rd</sup> year | 36 (26.3%) | 44 (28.9%) | 27 (38.0%) | 32 (26.7%) | 19 (22.9%) | 20 (24.1%) | 33 (26.6%) | 23 (25.8%) | 234<br>(27.2%) |
|  | 4<sup>th</sup> year | 27 (19.7%) | 34 (22.4%) | 7 (9.9%) | 40 (33.3%) | 25 (30.1%) | 17 (20.5%) | 31 (25.0%) | 25 (28.1%) | 206<br>(24.0%) |
| Owned a smartphone | No | 71 (51.8%) | 69 (45.4%) | 40 (56.3%) | 34 (28.3%) | 42 (50.6%) | 33 (39.8%) | 83 (66.9%) | 49 (55.1%) | 421<br>(49.0%) |
|  | Yes | 66 (48.2%) | 83 (54.6%) | 31 (43.7%) | 86 (71.7%) | 41 (49.4%) | 50 (60.2%) | 41 (33.1%) | 40 (44.9%) | 438<br>(51.0%) |
| Easy access to mobile data plans | No | 32 (48.5%) | 40 (48.2%) | 19 (61.3%) | 6 (7.0%) | 10 (24.4%) | 22 (44.0%) | 22 (53.7%) | 14 (35.0%) | 165<br>(37.7%) |
|  | Yes | 34 (51.5%) | 43 (51.8%) | 12 (38.7%) | 80 (93.0%) | 31 (75.6%) | 28 (56.0%) | 19 (46.3%) | 26 (65.0%) | 273<br>(62.3%) |
| Past MHA experience | No | 105 (75.7%) | 130 (85.5%) | 67 (94.4%) | 82 (68.3%) | 63 (76.8%) | 74 (89.2%) | 115 (92.7%) | 84 (94.4%) | 720<br>(83.8%) |
|  | Yes | 33 (24.3%) | 22 (14.5%) | 4 (5.6%) | 38 (31.7%) | 19 (23.2%) | 9 (10.8%) | 9 (7.3%) | 5 (5.6%) | 139<br>(16.2%) |
| Months of prior MHA use to learn about sexual health |  | 6.58±5.75 | 4.64±4.17 | 6.25±4.65 | 4.66±3.4 | 5.61±5.69 | 3.11±2.85 | 6.33±7.35 | 4.2±4.66 | 5.28±4.83 |

### 3.2. Descriptive statistics of mean scores, by school

Table 2 presents the mean scores by school to illustrate the contextual profile of the establishments. On a Likert scale from 1 to 5, the highest scores concerned BI, PE and EE, revealing a strong intention among young people in Bukavu to use MHAs for CSE, supported by their perceived usefulness and ease of use. SI, FC and PR came in at moderate levels.

**Table 2:** Mean scores on variables, by school.

| Variables | *n* | BI | PE | EE | SI | FC | PR |
| --- | --- | --- | --- | --- | --- | --- | --- |
| Institut de Bagira | 137 | 4.27 ± 0.49 | 4.25 ± 0.47 | 3.86 ± 0.80 | 3.75 ± 0.76 | 3.66 ± 0.81 | 3.35 ± 0.90 |
| Institut Bangu | 152 | 4.10 ± 0.77 | 4.13 ± 0.73 | 3.59 ± 0.93 | 3.63 ± 0.85 | 3.22 ± 0.90 | 3.10 ± 1.00 |
| Athénée d'Ibanda | 71 | 4.27 ± 0.68 | 4.26 ± 0.65 | 3.74 ± 0.87 | 3.33 ± 0.98 | 3.32 ± 0.94 | 3.18 ± 1.00 |
| Collège Alfajiri | 120 | 3.93 ± 0.82 | 3.93 ± 0.70 | 3.56 ± 0.84 | 3.22 ± 0.86 | 3.17 ± 0.82 | 3.50 ± 1.06 |
| Institut de Kadutu | 83 | 4.28 ± 0.53 | 4.27 ± 0.58 | 3.82 ± 0.66 | 4.06 ± 0.77 | 3.33 ± 0.83 | 3.33 ± 0.95 |
| Institut Kasali | 83 | 4.41 ± 0.68 | 4.42 ± 0.47 | 4.02 ± 0.79 | 3.67 ± 0.90 | 3.47 ± 0.76 | 3.31 ± 0.97 |
| Institut Avenir | 124 | 4.32 ± 0.45 | 4.11 ± 0.61 | 3.78 ± 0.71 | 3.77 ± 0.68 | 3.59 ± 0.83 | 3.42 ± 0.87 |
| Institut Imani Panzi | 89 | 4.26 ± 0.53 | 4.19 ± 0.61 | 3.70 ± 0.74 | 3.54 ± 0.81 | 3.42 ± 0.87 | 3.21 ± 0.85 |
| Total/Gen. avg. | 859 | 4.21 ± 0.65 | 4.18 ± 0.63 | 3.74 ± 0.81 | 3.62 ± 0.85 | 3.40 ± 0.86 | 3.30 ± 0.96 |

### 3.3. Predictors of intention of young people to use MHAs for CSE (H1 and H2): bivariate and multivariate analyses

Results regarding H1 and H2 are presented in Tables 3, 4, and 5.

**Table 3:**
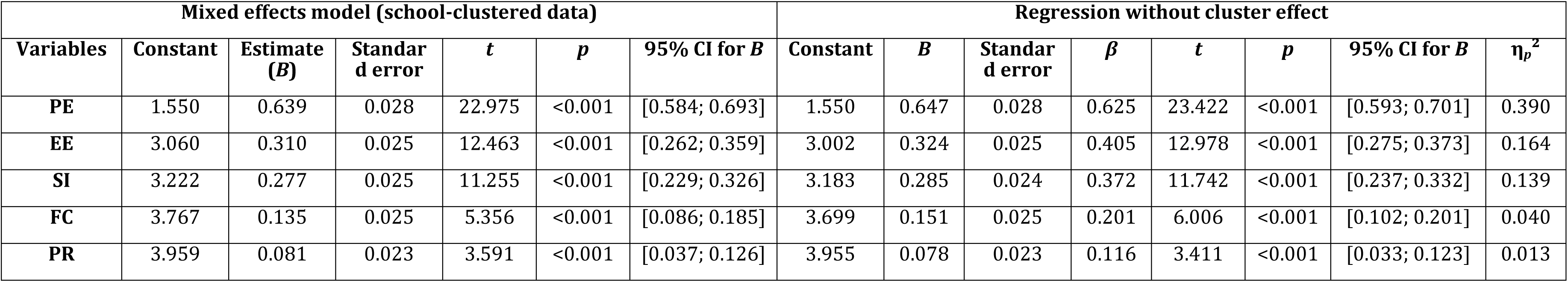
Relationships between PE, EE, SI, FC, PR and intention to use MHAs for CSE: H1 and H2 tests.

| Mixed effects model (school-clustered data) |  |  |  |  |  |  | Regression without cluster effect |  |  |  |  |  |  |  |
| --- | --- | --- | --- | --- | --- | --- | --- | --- | --- | --- | --- | --- | --- | --- |
| Variables | Constant | Estimate ($B$) | Standard error | $t$ | $p$ | 95% CI for $B$ | Constant | $B$ | Standard error | $\beta$ | $t$ | $p$ | 95% CI for $B$ | $\eta_p^2$ |
| PE | 1.550 | 0.639 | 0.028 | 22.975 | <0.001 | [0.584; 0.693] | 1.550 | 0.647 | 0.028 | 0.625 | 23.422 | <0.001 | [0.593; 0.701] | 0.390 |
| EE | 3.060 | 0.310 | 0.025 | 12.463 | <0.001 | [0.262; 0.359] | 3.002 | 0.324 | 0.025 | 0.405 | 12.978 | <0.001 | [0.275; 0.373] | 0.164 |
| SI | 3.222 | 0.277 | 0.025 | 11.255 | <0.001 | [0.229; 0.326] | 3.183 | 0.285 | 0.024 | 0.372 | 11.742 | <0.001 | [0.237; 0.332] | 0.139 |
| FC | 3.767 | 0.135 | 0.025 | 5.356 | <0.001 | [0.086; 0.185] | 3.699 | 0.151 | 0.025 | 0.201 | 6.006 | <0.001 | [0.102; 0.201] | 0.040 |
| PR | 3.959 | 0.081 | 0.023 | 3.591 | <0.001 | [0.037; 0.126] | 3.955 | 0.078 | 0.023 | 0.116 | 3.411 | <0.001 | [0.033; 0.123] | 0.013 |

**Table 4:**
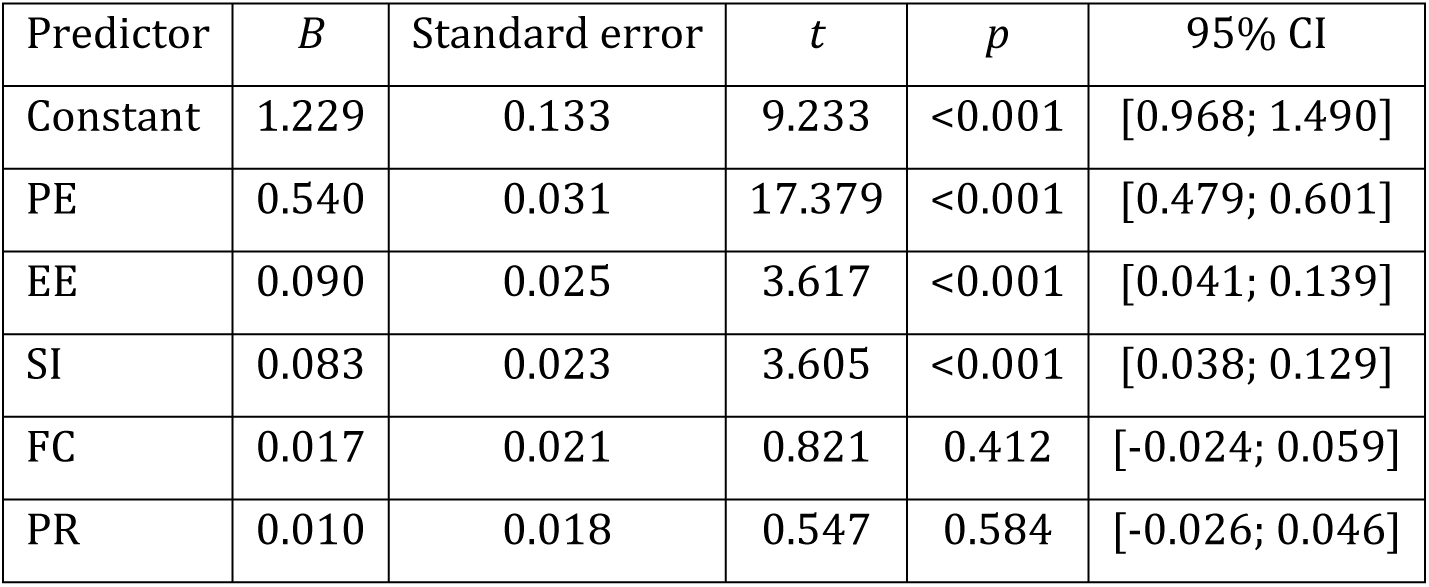
Mixed effects linear regression with adjustment for school clustering.

| Predictor | *B* | Standard error | *t* | *p* | 95% CI |
| --- | --- | --- | --- | --- | --- |
| Constant | 1.229 | 0.133 | 9.233 | <0.001 | [0.968; 1.490] |
| PE | 0.540 | 0.031 | 17.379 | <0.001 | [0.479; 0.601] |
| EE | 0.090 | 0.025 | 3.617 | <0.001 | [0.041; 0.139] |
| SI | 0.083 | 0.023 | 3.605 | <0.001 | [0.038; 0.129] |
| FC | 0.017 | 0.021 | 0.821 | 0.412 | [-0.024; 0.059] |
| PR | 0.010 | 0.018 | 0.547 | 0.584 | [-0.026; 0.046] |

**Table 5:** Multiple linear regression without cluster effect.

| Predictor | *B* | Standard error | $\beta$ | *t* | *p* | 95% CI for B | $\eta_p^2$ |
| --- | --- | --- | --- | --- | --- | --- | --- |
| Constant | 1.185 | 0.130 | — | 9.098 | <0.001 | [0.930; 1.441] | — |
| PE | 0.542 | 0.031 | 0.523 | 17.457 | <0.001 | [0.481; 0.603] | 0.263 |
| EE | 0.092 | 0.025 | 0.115 | 3.664 | <0.001 | [0.043; 0.141] | 0.015 |
| SI | 0.086 | 0.023 | 0.113 | 3.785 | <0.001 | [0.042; 0.131] | 0.017 |
| FC | 0.024 | 0.021 | 0.031 | 1.127 | 0.260 | [-0.018; 0.065] | 0.001 |
| PR | 0.009 | 0.018 | 0.013 | 0.481 | 0.631 | [-0.027; 0.045] | 0.000 |

#### 3.3.1. Bivariate analyses

Table 3 presents the results of the bivariate analyses under a mixed effects model with school-clustered data and the regression without the cluster effect.

Two complementary statistical approaches were used to assess the associations between PE, EE, SI, FC, PR and intention to use MHAs for CSE. First, simple regressions were run to estimate the bivariate relationships of each centred predictor and to examine individual links with intention. Then, mixed effects linear models were used with school-clustered data to control for intra-group dependence. The results of the two approaches converged and showed that PE, EE, SI and FC had a significant positive influence on intention to use MHAs for CSE among young people in Bukavu. On the other hand, PR had a significant effect, albeit a very weak one, that did not correspond to the negative effect expected on intention. The estimates yielded by the mixed model ranged from 0.639 (PE) to 0.081 (PR), all of which reached statistical significance (*p* < 0.001) and had a 95% CI that excluded zero. The simple regressions revealed a hierarchy: PE (β = 0.625) > EE (β = 0.405) > SI (β = 0.372) > FC (β = 0.201) > PR (β = 0.116). An analysis of explained variance (η*_p_*²) showed that PE accounted for 39% of the variance, compared with 16.4% for EE, 13.9% for SI, 4% for FC, and 1.3% for PR.

#### 3.3.2. Multivariate analyses

Table 4 presents the results of the mixed effects linear regression analyses with school-clustered data to adjust for intra-group dependence. These analyses showed that PE, EE and SI were positively and significantly associated with intention to use MHAs for CSE (*p* < 0.001). On the other hand, FC and PR did not prove significantly associated with intention to use.

Table 5 shows that, in the multiple linear regression model, PE, EE and SI remained the three statistically significant predictors of intention to use MHAs for CSE. PE (*β* = 0.523; *p* < 0.001) remained the strongest predictor, alone accounting for 26.3% of the variance in intention. While EE (*β* = 0.115; *p* < 0.001), and SI (*β* = 0.113; *p* < 0.001) were also significant, their explanatory contribution was modest. FC and PP did not prove significant, suggesting that their effect diminished when other factors were controlled for. These results supported HI in part and nullified H2.

### 3.4. Analysis of moderating effects of age, gender, and past MHA experience on digital CSE use of young Congolese: H3 to H5

H3 to H5 postulated that certain sociodemographic characteristics moderated the relationships between PE, EE, SI, FC, PR and intention to use MHAs for CSE among young Congolese. H3 stipulated that age modulated these relations and that the effect was differentiated by age; H4, that gender moderated these relationships; and H5, that past MHA experience had a positive moderating effect.

Analysis of the interaction effects showed that age had a significant moderating effect on the relationships between each of the predictors (PE, EE, SI, FC, PR) and intention to use, with the effects ranging from *F*(1, 849–854) = 9.97 to 20.82 and *p*–values ranging from <0.001 to 0.002 (Table 6). These results supported H3, according to which age moderated the effects of the predictors on intention to use MHAs for CSE. The younger students demonstrated a greater sensitivity to PE, EE, and SI, with the moderating effect of age slightly more pronounced for SI. For FC and PR, the differences between younger and older students remained statistically significant but associated with smaller effect sizes.

**Table 6:** Moderating effects of age, gender, and past MHA experience on intention to use digital CSE.

| Predictors | Moderators | *df*<br>numerator | *df*<br>denominator | *F* | *p* | $\eta_p^2$ |
| --- | --- | --- | --- | --- | --- | --- |
| **PE** | Age | 1 | 854.048 | 15.663 | <0.001 | 0.019 |
|  | Gender | 1 | 853.343 | 4.299 | 0.038 | 0.006 |
|  | Experience | 1 | 849.622 | 3.126 | 0.077 | 0.004 |
| **EE** | Age | 1 | 854.284 | 19.482 | <0.001 | 0.024 |
|  | Gender | 1 | 853.887 | 2.998 | 0.084 | 0.003 |
|  | Experience | 1 | 852.310 | 2.165 | 0.142 | 0.002 |
| **SI** | Age | 1 | 853.191 | 20.815 | <0.001 | 0.026 |
|  | Gender | 1 | 851.617 | 0.117 | 0.733 | 0.000 |
|  | Experience | 1 | 847.738 | 7.377 | 0.007 | 0.008 |
| **FC** | Age | 1 | 851.871 | 13.517 | <0.001 | 0.017 |
|  | Gender | 1 | 851.501 | 1.455 | 0.228 | 0.001 |
|  | Experience | 1 | 849.383 | 1.066 | 0.302 | 0.001 |
| **PR** | Age | 1 | 849.272 | 9.973 | 0.002 | 0.011 |
|  | Gender | 1 | 851.273 | 0.316 | 0.574 | 0.000 |
|  | Experience | 1 | 847.959 | 0.154 | 0.694 | 0.000 |

Regarding gender, only its moderating effect on the relationship between PE and BI proved statistically significant (*F* (1, 853) = 4.30; *p* = 0.038; η*_p_*² = 0.006), indicating that PE influenced boys and girls differently. On the other hand, its interactions with the other predictors were not significant. These results supported in part H4, according to which gender moderated the relationships between PE, EE, SI, and PR with intention to use MHAs for CSE among young people in Bukavu.

As for past MHA experience, only the SI x BI interaction proved significant (*F* (1, 848) = 7.38; *p* = 0.007; η*_p_*² = 0.008), suggesting a more pronounced social support effect among participants already familiar with these tools. These results supported H5 only in part, as past MHA experience had a significant moderating effect only on the effect of SI among the five predictors tested.

Figures 3, 4, and 5 present the estimates for BI by level of centred predictor (age, PE, and SI).

**Fig 3:**
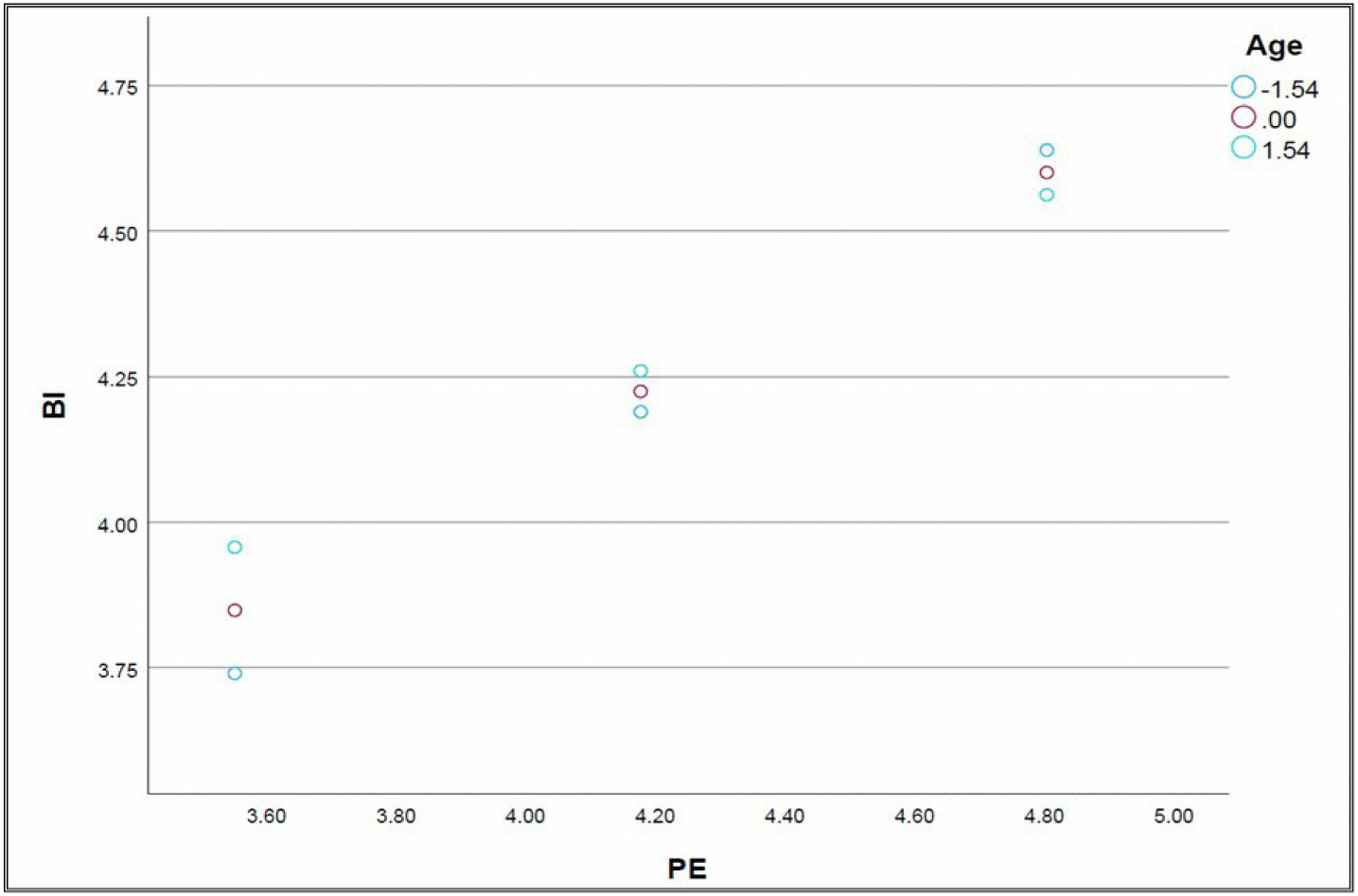
Centred PE x Centred Age interaction effect on intention to use CSE MHAs

**Fig 4:**
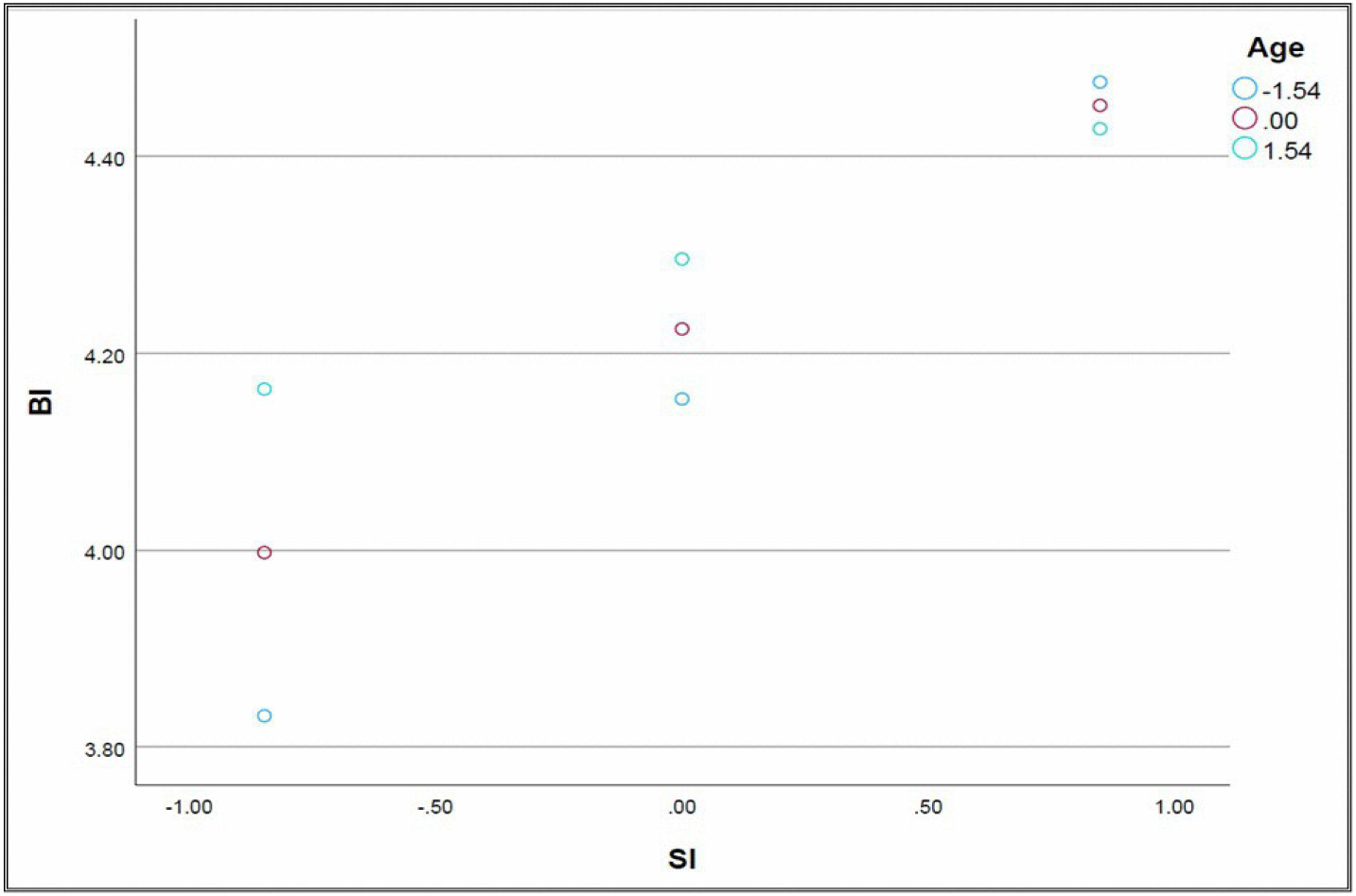
SI x Centred Age interaction effect on intention to use CSE MHAs

**Fig 5:**
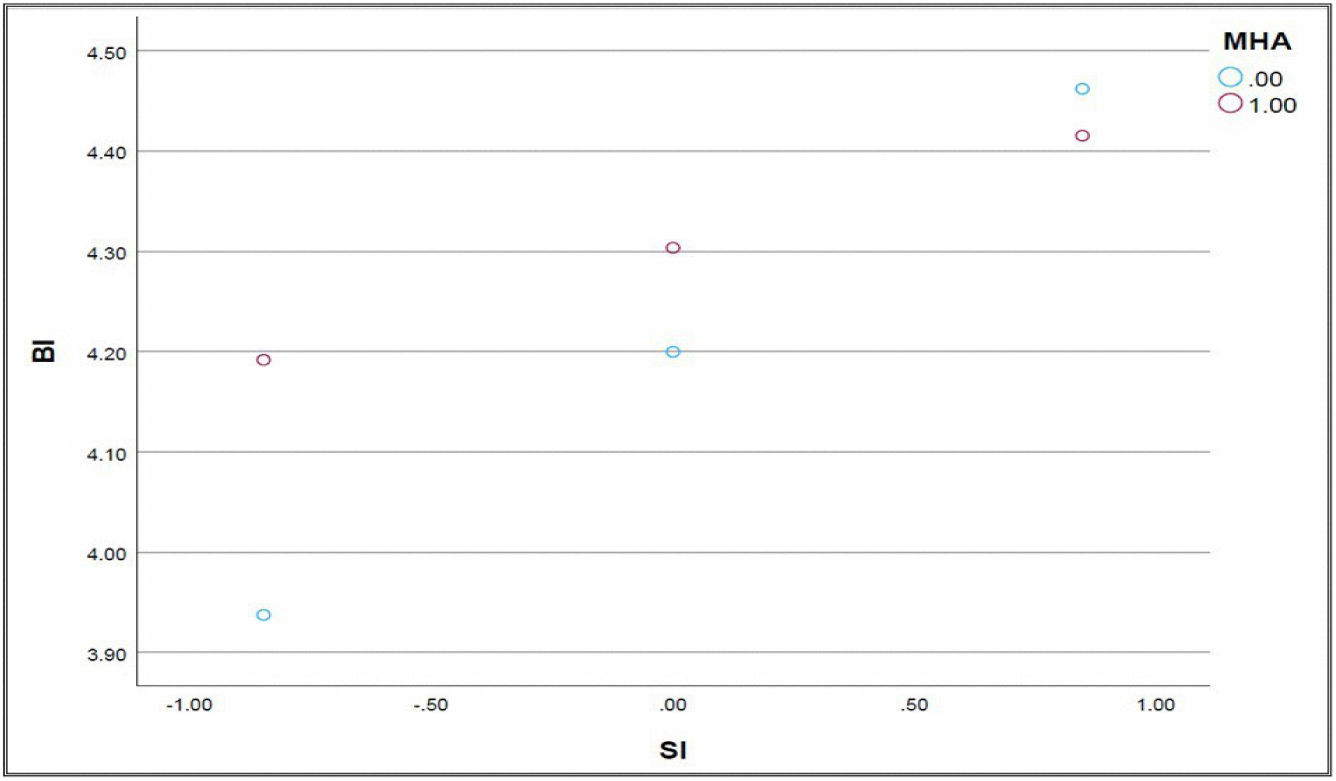
Moderating effect of past MHA experience on relationship between SI and intention to use CSE MHAs

Figure 5 illustrates the effects of the Experience x SI interaction. It shows that BI increased along with SI, both among participants who had already used applications of this type and among those with no past experience. However, the differences in intention between these two groups remained relatively modest, which was consistent with a weaker moderating effect of past MHA experience. In sum, these figures indicate that age played a more important role than did past MHA experience in how young people responded to PE and SI.

### 3.5. Final model testing H1 to H5 regarding intention among young Congolese to use CSE MHAs

The hierarchical regression model allowed us to test H1 to H5 in an integrated manner. The first model (Table 7) included the direct effects of the main predictors (PE, EE, SI, FC, PR) and of the centred moderators (age, gender, experience). The second model (Table 8) included also the moderating interactions to refine the explanation of BI. In Table 7, *R*² indicates the portion of the variance accounted for by the successive models. Model 1 presented an *R*² of 0.424, indicating that 42.4% of the variance was accounted for by the main predictors (*p* < 0.001). Adding the interactions in Model 2 improved explanatory power slightly but significantly (*R*² = 0.441; Δ*R*² = 0.017; *p* < 0.001).

**Table 7:** Stepwise hierarchical regression: coefficients of determination and improved explanatory power.

| Model | $R$ | $R^2$ | Adjusted $R^2$ | Standard error of estimate | $\Delta R^2$ | $\Delta F$ | $df1$ | $df2$ | $p (\Delta F)$ |
| --- | --- | --- | --- | --- | --- | --- | --- | --- | --- |
| 1 | 0.651 | 0.424 | 0.419 | 0.495 | 0.424 | 89.099 | 7 | 849 | <0.001 |
| 2 | 0.664 | 0.441 | 0.432 | 0.489 | 0.017 | 4.324 | 6 | 843 | <0.001 |

**Table 8:**
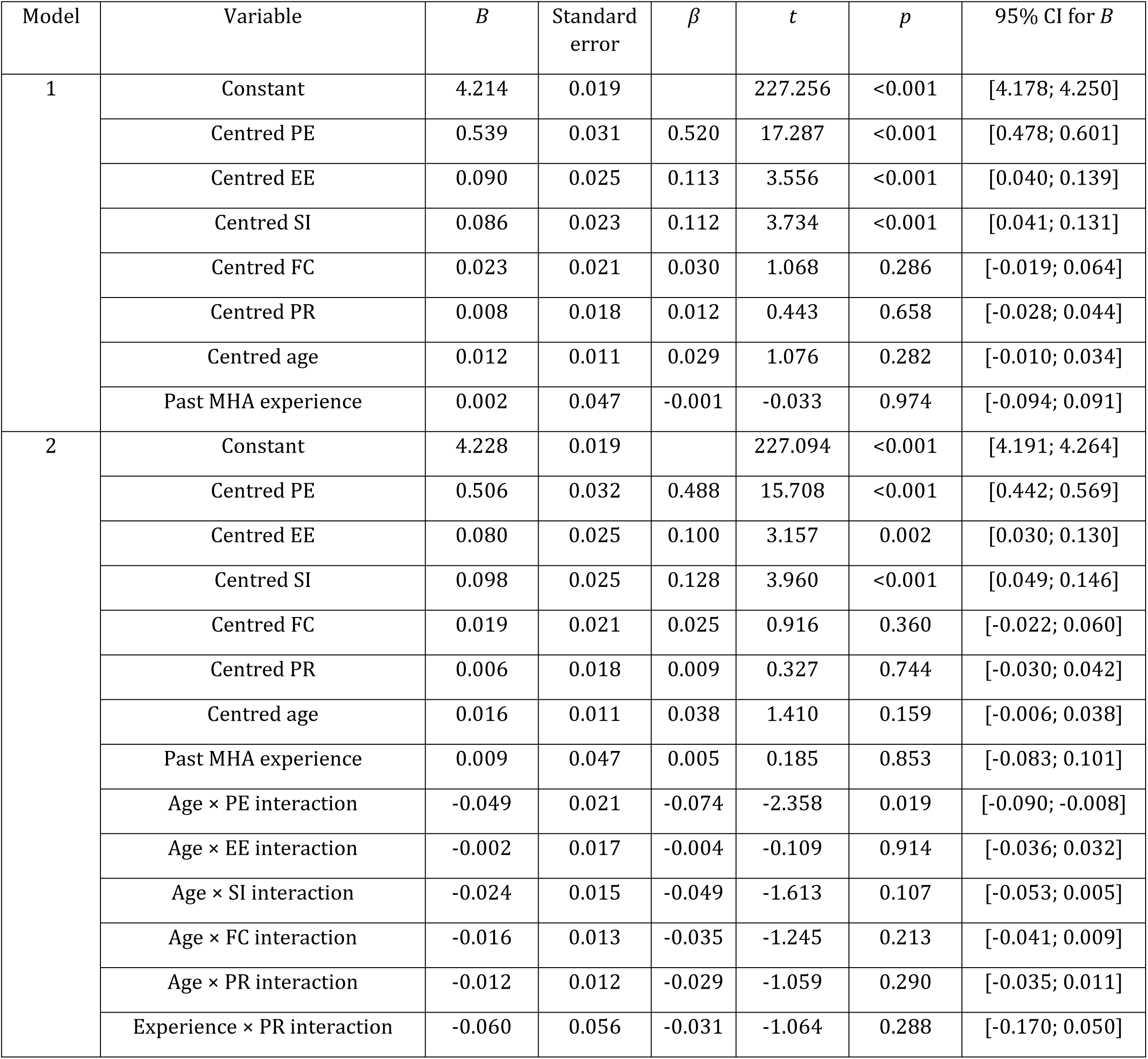
Final hierarchical regression model for predictors of BI.

| Model | Variable | *B* | Standard error | $\beta$ | *t* | *p* | 95% CI for *B* |
| --- | --- | --- | --- | --- | --- | --- | --- |
| 1 | Constant | 4.214 | 0.019 |  | 227.256 | <0.001 | [4.178; 4.250] |
|  | Centred PE | 0.539 | 0.031 | 0.520 | 17.287 | <0.001 | [0.478; 0.601] |
|  | Centred EE | 0.090 | 0.025 | 0.113 | 3.556 | <0.001 | [0.040; 0.139] |
|  | Centred SI | 0.086 | 0.023 | 0.112 | 3.734 | <0.001 | [0.041; 0.131] |
|  | Centred FC | 0.023 | 0.021 | 0.030 | 1.068 | 0.286 | [-0.019; 0.064] |
|  | Centred PR | 0.008 | 0.018 | 0.012 | 0.443 | 0.658 | [-0.028; 0.044] |
|  | Centred age | 0.012 | 0.011 | 0.029 | 1.076 | 0.282 | [-0.010; 0.034] |
|  | Past MHA experience | 0.002 | 0.047 | -0.001 | -0.033 | 0.974 | [-0.094; 0.091] |
| 2 | Constant | 4.228 | 0.019 |  | 227.094 | <0.001 | [4.191; 4.264] |
|  | Centred PE | 0.506 | 0.032 | 0.488 | 15.708 | <0.001 | [0.442; 0.569] |
|  | Centred EE | 0.080 | 0.025 | 0.100 | 3.157 | 0.002 | [0.030; 0.130] |
|  | Centred SI | 0.098 | 0.025 | 0.128 | 3.960 | <0.001 | [0.049; 0.146] |
|  | Centred FC | 0.019 | 0.021 | 0.025 | 0.916 | 0.360 | [-0.022; 0.060] |
|  | Centred PR | 0.006 | 0.018 | 0.009 | 0.327 | 0.744 | [-0.030; 0.042] |
|  | Centred age | 0.016 | 0.011 | 0.038 | 1.410 | 0.159 | [-0.006; 0.038] |
|  | Past MHA experience | 0.009 | 0.047 | 0.005 | 0.185 | 0.853 | [-0.083; 0.101] |
|  | Age × PE interaction | -0.049 | 0.021 | -0.074 | -2.358 | 0.019 | [-0.090; -0.008] |
|  | Age × EE interaction | -0.002 | 0.017 | -0.004 | -0.109 | 0.914 | [-0.036; 0.032] |
|  | Age × SI interaction | -0.024 | 0.015 | -0.049 | -1.613 | 0.107 | [-0.053; 0.005] |
|  | Age × FC interaction | -0.016 | 0.013 | -0.035 | -1.245 | 0.213 | [-0.041; 0.009] |
|  | Age × PR interaction | -0.012 | 0.012 | -0.029 | -1.059 | 0.290 | [-0.035; 0.011] |
|  | Experience × PR interaction | -0.060 | 0.056 | -0.031 | -1.064 | 0.288 | [-0.170; 0.050] |

Table 8 presents the coefficients for the main predictors and the moderating interactions. The final model confirmed the direct effects of PE (*β* = 0.488; *p* < 0.001), EE (*β* = 0.100; *p* = 0.002), and SI (*β* = 0.128; *p* < 0.001) on BI to use MHAs for CSE. Of the moderating effects tested, only the Age x PE interaction proved significant (*β* = –0.074; *p* = 0.019), indicating that PE had a more pronounced effect among younger students. No moderating effect was noted for gender and past MHA experience. These results supported H3 in full, nullified H4, and confirmed H5 in part.

## 4. DISCUSSION

Our study sought to identify the predictors of intention among young people in the DRC to use MHAs for CSE, based on the UTAUT2 extended to PR. The principal results indicated that PE, EE, and SI had a significant positive influence, whereas FC and PR had no notable effect. Age modulated all of the relationships tested, reflecting a greater sensitivity to perceived usefulness, ease of use, SI, as well as FC and PR, and underscoring the appeal of early e-interventions for prevention, CSE, and the adoption of responsible sexual health behaviours. The final model accounted for 44% of the variance, which attests to solid explanatory power.

In our study, the explained variance (*R*² = 0.441) came in at an acceptable level. Generally speaking, technology acceptance research shows that models have explained from 17% to 53% of the variance in intention to use in adult or organizational samples, but that the UTAUT has often surpassed previous models by explaining as much as about 70% (49,52). From this perspective, 44% explained variance in a younger and more educated sample appears a solid result. Our results indicate that the individual dimensions of the UTAUT2 retain strong explanatory power at an early stage of the life course, including in the Congolese context marked by structural constraints. These values confirm, according to Ozili (71), that a coefficient of determination between 0.10 and 0.50 remains acceptable in the social sciences, especially when various explanatory variables prove significant, as was the case in our study. The model thus presents a consistent explanatory power: it supports H1, nullifies H2 and H4, confirms H3, and confirms H5 only in part.

Our first hypothesis postulated that PE, EE, SI, and FC had a positive influence on the intention of young people in the DRC to use MHAs for CSE. The results confirmed this part: PE, EE, and SI proved significant positive predictors. These findings are in line with those reported by Asadollahi et al. (51), which showed that these three factors had a positive influence on the intention of young pregnant Iranian women to use MHAs to access pre-natal care but that FC had no direct effect. They are in line also with the findings of a study that showed that these same factors were associated with the intention of young Kenyans, Nigerians and South Africans to access and interact with reliable sexuality information on the social networks (44). These convergences evidence the central role of these factors as predictors of intention to use mobile health technology, whereas the absence of an FC effect seems related more to context than to a theoretical limitation. This absence raises questions about the actual impact of FC in environments such as the city of Bukavu, and more widely the DRC, where digital infrastructures and technical support are emerging but still wanting.

Taken separately, each one of the three significant predictors in the final model has a specific influence on digital CSE acceptance. In our study, PE stood out as the most consistent and influential predictor of intention to use MHAs for CSE. It remained significant even when analyzed jointly with the other factors and was moderated by participant age. This result is congruent with previous studies that identified PE as the principal predictor of mobile health intervention acceptance (72). Modulated by age (19,36), PE stimulates motivation and engagement among young people, thus bolstering the effectiveness of the interventions. In other words, the more that young people perceive interventions to have a practical usefulness, the more that young people invest in the interventions. Positive expectations thus constitute a key lever of behavioural change by fostering engagement and practical benefits (72). In the context of digital CSE in the DRC, a strong PE might boost motivation among young people to translate knowledge into responsible health behaviours, particularly among those more inclined to accept technology perceived as effective, esteem-building, and gratifying (36,41,42). It is therefore essential to reinforce this dimension through motivational messages, constructive feedback, and the concrete demonstration of expected benefits—be it faster access to CSE or improved health— especially in a context where individual success and performance are valorized (73).

Aside from PE, EE stood out as well in the final model. It, too, was moderated by age: Younger students more familiar with digital environments had a higher appreciation for the simplicity and intuitiveness of MHAs. Similar results were observed in the young urban populations in Turkey and Japan. In Turkey, perceived ease of use—one of the four items measuring EE (49)—was found to have a significant influence on intention to use MHAs (74). In Japan, the perception of an intuitive, user-friendly interface, too, was found to reinforce intention among young adults (54), underscoring the importance of ease of use in MHA acceptance. These findings from studies conducted in technologically advanced Asian societies suggest that EE is shaped also by sociocultural dimensions. A recent meta-analysis involving 6,128 participants including young people (75) showed that EE was positively modulated by the local culture, particularly collectivism, femininity, and masculinity. Collectivism valorizes social cohesion and interpersonal relationships. Femininity places more of an emphasis on quality of life, gender equality, security, and collective well-being. Masculinity, however, prioritizes competitiveness, personal success, and material rewards and is associated with weak avoidance of uncertainty and faster adoption of technological innovations, especially mobile devices (73,75). According to Hofstede et al. (73), the masculinity index, normalized to 100 points, varies from 0 for very feminine societies to 100 for the most masculine. The countries of sub-Saharan Africa, such as the DRC, score in the range of 40 to 63 (76), which translates a moderate masculine orientation. In these cultures, Zhang et al. (75) showed that individuals ascribe more importance to perceived ease of use. This cultural disposition could thus facilitate the receptiveness of young Congolese to digital CSE, which is perceived as a useful technological innovation both socially valorized and beneficial to the individual. This innovation has become a reality through MHAs, which today have become an ideal means of offering young people CSE tailored to their needs. Designed for young people but especially co-designed with them, these applications should be simple, intuitive, user-friendly, offline-first, and endowed with an attractive design, in order to reinforce their cultural acceptability (73,77). They should also allow rapid onboarding and progressive ramp-up to facilitate learning and self-learning by young people. Such an ergonomic and participatory approach is believed to foster not only acceptance, but also user loyalty and sustained app use.

In addition to PE and EE, a third predictor proved significant in our final model: SI. Its effect was modulated by age: It was stronger among users already familiar with these apps, for whom perceived social support acted as an SI lever and boosted their intention to use these tools. These results contrast with results observed in Germany, characterized by an individualist culture (73,75), where SI was found to have no significant effect on the intention of 105 young people to use an educational MHA for prevention in high-risk groups (37). In such a context, autonomy, independence, and personal responsibility override collective expectations. Our results differed, also, from those reported in a study conducted in Kenya (45), where SI was found to have no significant effect on the intention of 77 young users of a digital sexuality education tool. Yet, with a score of 25 out of 100 for individualism (76), Kenya is a country with a collectivist culture that valorizes interpersonal relationships, where the influence of peers and persons deemed important should play a determining role in the acceptance of CSE MHAs. On the other hand, our results corroborate those of Akiogbe et al. (78), whose comparative analysis of Japanese and Chinese young people, both from collectivist cultures (55,73), showed that cultural norms and social dynamics had a strong influence on mobile health technology acceptance. These cultural contrasts are particularly illuminating: they show that intention to use technology is not a universal variable but a contextual one shaped by the values, norms and social dynamics specific to each society. In the DRC, where social ties remain strong, the influence of parents, peers, teachers, and community leaders merits special attention. From this point of view, it is essential that the development and implementation of CSE MHAs reflect the local sociocultural dynamics. This supposes integrating social support, community forums and e-mentoring functions (79) in order to bolster the trust, sense of social valorization, and engagement of young people towards these digital health education tools.

Unlike the three previous predictors, FC and PR did not have a significant effect in the final model. Regarding FC, our results differ from those of previous studies where the young participants had past MHA experience (74,80). In our study, only 16.2% of the participants had already been using an MHA to learn about sexual health, on average for about five months (5.28 ± 4.83). This poor familiarity with MHAs might explain in part the lack of statistical significance observed. The students did not yet have the resources, technological support, and practical knowledge required to use apps for the purpose of CSE, all the more so that only 51% owned a smartphone. Still, in the Congolese context marked by digital inequalities and limited institutional support (47,48), FC remains crucial. FC can be reinforced through computer skills workshops, digital health training sessions, technical local mentoring, and offline user-friendly environments, in order to consolidate trust, autonomy and sustained use of digital CSE. However, even a facilitating environment might not suffice if perceptions related to security and confidentiality are not taken into account. This is why our second hypothesis concerned the role of PR in intention to use. As it turns out, this factor did not have a significant effect, thus nullifying H2, according to which PR would have a negative influence on intention to use MHAs. This result is consistent with those of studies carried out in Poland (39), Germany (56), and Japan (54), where confidentiality and security concerns did not damper intention to use. Among young people, this absence of effect is explained by a high level of trust towards MHAs and the search for autonomy and effectiveness (39,54). In the Congolese context, this tendency could be amplified by the priority granted to PE, EE, and SI, over the protection of personal data. Rapid access to useful and reliable information, interactivity, and sense of autonomy seem, as a result, to trump security concerns, thus attenuating the effect of PR on intention to use CSE MHAs (54,81). Concerns regarding confidentiality and cyber-threats appear to take a back seat to the search for effectiveness and social recognition. PR remains latent, but not a priority, in the intention to use digital CSE among young Congolese. However, this weak sensitivity to risk underscores the necessity of reinforcing awareness of digital security and the protection of personal data in CSE programs to encourage young people to use MHA responsibly and trustfully.

Overall, these results provide a solid empirical basis for designing mobile health interventions that are culturally grounded, user centric, and adapted to Francophone contexts with limited resources, including to promote digital CSE to young people in Francophone sub-Saharan Africa.

### Strengths and limitations

Our study has numerous strengths worth noting. It rests on a large, stratified, multi-site random sample recruited in public confessional and non-confessional schools in urban and suburban centres, thus reinforcing the contextual richness of the data. Adapting the UTAUT2 model by incorporating PR constituted a contextual adjustment aimed at strengthening the model’s relevance in the study of intention to use digital CSE. The use of a UTAUT questionnaire validated in a Francophone context, the detailed description of the school system in the DRC, and the use of analyses adjusted to school-clustered student data to control for unmeasured institutional heterogeneity support the robustness of the estimates at the individual level. However, our study also has limitations that need to be pointed out. The Cronbach’s alpha values observed for some subscales—SI (α = 0.593), FC (α = 0.539), and PR (α = 0.563)—indicate low internal consistency, though it is deemed acceptable for scales comprising a small number of items. The observational nature of the study, marked by the absence of the manipulation and control of the variables, does not allow inferring any causality. Moreover, our analysis, which was centred on the individual characteristics of the students and did not take account of organizational variables, did not allow us to identify the institutional factors that might explain differences across schools. Furthermore, the sample, which was restricted to the city of Bukavu, reflected a particular academic and digital context. Despite these limitations, our results are generalizable to young students in Bukavu and can shed light on Congolese or African contexts with similar characteristics.

### Practice and research implications

This study makes several important practical and scientific contributions. It fills a gap in the literature on intention to use mobile health technologies in the DRC and Francophone sub-Saharan Africa. Based on a targeted operationalization of the UTAUT2, extended to include PR, this study sheds light on the factors that influence intention to use CSE MHAs among young people. Regarding practice, the results could guide decision-makers, community stakeholders, and app designers towards tailored solutions and develop simple, intuitive and reliable tools grounded in the local culture and integrated in education and health policies, to help young people become autonomous and to foster their sustained engagement. At the scientific level, our study opens the door to qualitative research on digital CSE acceptance. It also highlights the need to analyze the effective use of MHAs beyond intention to use in order to better understand how intention translates into actual behaviour in the Congolese educational and community contexts. Lastly, it points to why it would be interesting next to employ longitudinal and multi-level approaches to examine how this intention changes over time and how school-related contextual factors influence the acceptance and use of digital CSE. The differences observed on the bases of age and SI underscore the need for nursing interventions grounded in proximity, solidarity, and respect for human dignity to provide support tailored to the realities of young people. In African contexts, such an approach meshes logically with notions of collective responsibility and interdependence consistent with the principles of the Ubuntu philosophy. Finally, our results urge us to explore, through MHAs, how some essential components of CSE—still alarming in the DRC—are offered to young people, notably birth-control services and safe abortion care, pre- and post-natal care, as well as prevention and care relative to HIV infection and gender-based violence.

## Conclusion

Based on the UTAUT2, our study allowed us to identify the principal predictors of intention to use MHAs for CSE among young people in the DRC. In sum, performance expectancy, effort expectancy, and social influence were found to have a positive influence, whereas facilitating conditions and perceived risk proved to have no significant effect. Age modulated all of the relationships tested by strengthening the effect of the UTAUT2 predictors among young people, reflecting a greater sensitivity to perceived usefulness, ease of use, and social influence at this stage of the life course, whereas gender and past MHA experience only had a more targeted moderating effect. These results speak to the willingness of young people to use simple, reliable, and socially valorizing digital tools, in a culture marked by solidarity and an openness to innovation. In this regard, the results suggest that their intention to use digital CSE fits in with a relational and community dynamic, consistent with the principles of the African philosophy of Ubuntu applied to innovation in the field of health. At the scientific and societal levels, our study adds to our understanding of the predictors of intention to use mobile health technologies in resource-limited Francophone contexts. It provides an empirical foundation for designing culturally grounded interventions aimed at reinforcing the sustainable use of digital CSE. Finally, our study opens up promising avenues for e-health in Francophone sub-Saharan Africa by contributing to promoting the health and well-being of young people and to meeting sustainable development goals.

## Data Availability

The data underlying the results presented in this study are available from the corresponding author upon reasonable request, subject to approval by the relevant ethics committees due to ethical restrictions.

## Acknowledgements

We would like to express our sincerest gratitude to the Sud-Kivu Provincial Principal Inspection Office of the Ministry of National Education and New Citizenship of the Democratic Republic of Congo, Sud-Kivu province, for its institutional support throughout this study. We would like to thank, also, the principals of the schools that participated in the study and the focal point teachers for their collaboration, availability and engagement in coordinating the data collection. Our sincerest thanks go out, also, to the students who took part in the study. Their availability, trust, and enthusiasm in completing our questionnaire were essential to this research.

## Funding sources

This study received financial support from the Research Chair in Innovative Nursing Practices of the Nursing Faculty of the Université de Montréal. It was supported, also, by the Canadian Behavioural Interventions and Trials Network (CBITN), which receives funding from the Canadian Institutes of Health Research (CIHR) (Grant reference number: CTT-184895).

## Competing interest

The authors have no conflict of interest to declare.

## Author contribution

**FKM:** Conceptualization, data curation, funding acquisition, investigation, methodology, project administration, resources, visualization, writing – original draft preparation

**JC**: Conceptualization, supervision, validation, writing – review & editing

**AB**: Writing – review & editing

**CA**: Writing – review & editing

**MC**: Formal analysis, writing – review & editing

**MH**: Conceptualization, supervision, validation, writing – review & editing.

## Abbreviations and symbols

*B*: Unstandardized coefficient
BI: Behavioural intention
CSE: Comprehensive sexuality education
DRC: Democratic Republic of the Congo
EE: Effort expectancy
H1: First research hypothesis
H2: Second research hypothesis
H3: Third research hypothesis
H4: Fourth research hypothesis
H5: Fifth research hypothesis
HM: Hedonic motivation
MHA: Mobile health application
PE: Performance expectancy
PR: Perceived risk
PV: Price value
SI: Social influence
STROBE: Strengthening the Reporting of Observational Studies in Epidemiology
TAM: Technology Acceptance Model
UTAUT: Unified Theory of Acceptance and Use of Technology
UTAUT2: Extended Unified Theory of Acceptance and Use of Technology
β: Standardized coefficient
η*_p_*²: Partial eta squared
*R*²: Coefficient of determination

## Notes

### Competing Interest Statement

The authors have declared no competing interest.

### Funding Statement

Yes

### Author Declarations

The study was approved by the Université de Montréal Health and Science Research Ethics Committee (CERSSES 2024-6039) and by the DRC National Health Ethics Committee (CNES 001/DPSK/222PM/2024). Participation was voluntary, and informed consent was obtained from all participants. For participants aged 18 years and older, written informed consent was obtained directly. For minors aged 15 to 17 years, consent was obtained from school principals acting as institutional legal guardians, and assent was obtained from the students prior to data collection. No personal identifiers were collected.

